# AI-Driven Early Detection of Severe Influenza in Jiangsu, China: A Deep Learning Model Validated Through The Design of Multi-Center Clinical Trials and Prospective Real-World Deployment

**DOI:** 10.1101/2025.02.12.25322194

**Authors:** Yifei Chen, Yan Bo

## Abstract

Influenza causes about 650,000 deaths worldwide each year, and the high mortality rate of severe cases is closely related to subjective bias in clinical assessment and inconsistent diagnostic and treatment standards. To this end, this study developed and validated a deep learning-based model for early diagnosis of severe influenza that optimises risk stratification by integrating clinical data from multiple sources. The study included 87 tertiary general hospitals in Jiangsu Province, China, and used a five-stage validation framework (model development, external validation, multi-reader study, randomised controlled trial, and prospective validation) to analyse electronic health record data covering demographic, symptomatic, laboratory indicators, and imaging features between 2019 and 2025. Expected results showed that the model-assisted diagnosis had a significantly higher AUC value of 0.18 (95% CI: 0.14-0.22) and a 32% lower rate of misdiagnosis compared to traditional clinical assessment, and performed consistently in elderly and chronically ill patients and in hospitals in resource-limited areas (subgroups with AUCs of >0.82 in all cases). The expectation of the study will be realised that the model can effectively improve the early recognition of severe influenza by dynamically integrating multidimensional information, especially for scenarios where healthcare resources are unevenly distributed. The implementation of this study followed strict ethical norms (JD-LK-2019-106-01, The Second Affiliated Hospital of Soochow University; 2024-10-02, The Affiliated Hospital of Yangzhou University), and the de-identified data were managed through an encrypted platform (osf.io/ayj75) and planned to open-source the model code in order to promote clinical translation and cross-region collaboration, and to provide a scalable influenza precision prevention and control decision support tools.

**Trial:** ChiCTR2000028883

**Registration DOI:** https://doi.org/10.17605/OSF.IO/SC93Y

## 1 Introduction

Influenza remains a global health threat, causing up to 650,000 deaths annually with a mortality rate of approximately 20%, particularly concentrated among high-risk populations such as infants, pregnant women, the elderly, and immunocompromised individuals (1,2). Despite advancements in predictive tools and antiviral therapies, mortality rates for severe influenza have not significantly declined over the past decade (3). A critical barrier lies in the persistent gap between clinical decision-making and timely risk stratification. Recent evidence highlights that physician overconfidence in subjective assessments and inconsistent definitions of “cure” within healthcare systems contribute to delayed interventions and misdiagnosis (4,5). For instance, Valenzuela-Sánchez et al. (2024) reported that 34% of severe influenza cases were initially misclassified as mild due to overreliance on nonspecific symptom evaluation (6). This underscores the urgent need for objective, data-driven tools to augment clinical judgment.

Artificial intelligence (AI), particularly deep learning, has demonstrated transformative potential in medical diagnostics by integrating heterogeneous data (e.g., clinical symptoms, biomarkers, imaging) to identify high-risk patients (7,8). Recent studies, such as Yang et al. (2023), developed an AI model for influenza prognosis using urban-scale multisource data, achieving an AUC of 0.89 in retrospective cohorts (9). However, existing models often lack external validation across diverse healthcare settings or fail to address real-world clinical workflows (10). For example, Dai et al. (2024) emphasized that most AI diagnostic tools exhibit performance degradation when applied prospectively due to population heterogeneity and data drift (11). Furthermore, few studies have specifically targeted dynamic risk prediction for high-risk subgroups under real-time clinical constraints (12).

This study aims to bridge these gaps by developing and validating a deep learning-based diagnostic model for early identification of severe influenza through a multi-phase, multi-center trial. Our approach innovatively combines:

1. Multi-source data integration: Clinical, laboratory, and demographic variables tailored to high-risk populations.
2. Rigorous validation framework: Five-phase evaluation spanning internal validation, comparative studies, randomized trials, and prospective real-world testing.
3. Clinician-AI collaboration: Assessing how model-assisted decisions impact diagnostic accuracy across physicians with varying experience levels.

We hypothesize that this model will significantly improve AUC (Δ >0.15) compared to physician-only assessments, reduce misdiagnosis rates by ≥30%, and maintain robustness across hospitals with varying resource levels. By addressing both algorithmic and human factors, this research seeks to establish a scalable solution for mitigating influenza-related mortality, particularly in resource-constrained settings.

## 2 Methods

### 2.1 Study setting

This multi-center, multi-phase clinical trial is designed to develop and validate a deep learning-based diagnostic model for early identification of severe influenza. The study will be conducted across 87 tertiary general hospitals in all prefecture-level cities of Jiangsu Province, China. This study is a continuation of the 2019 investigation of influenza A (ChiCTR2000028883) and is registered on the OSF registry platform (13). This clinical trial will be reported according to SPIRIT 2013 standard specifications (14).

#### 2.1.1 Hospital Inclusion Criteria

Participating hospitals were required to meet stringent eligibility criteria to ensure data quality and comparability: all institutions must be government-certified tertiary general hospitals offering comprehensive emergency and inpatient services, with a documented annual caseload of ≥100 laboratory-confirmed influenza cases (via nucleic acid/antigen testing) over the preceding three years (2021–2023). Additionally, hospitals were mandated to maintain standardized electronic health records (EHRs) with structured fields for symptom documentation, laboratory results, imaging reports, and treatment protocols, enabling seamless data aggregation and analysis. Ethical compliance was ensured through institutional review board (IRB) approval and demonstrated capacity for longitudinal, anonymized data sharing. These criteria collectively guaranteed a robust, representative dataset reflective of diverse clinical practices and resource settings.

#### 2.1.2 Rationale for Multi-Center Design

The inclusion of hospitals across Jiangsu Province ensures generalizability by validating the model’s performance in diverse clinical environments, spanning urban centers with advanced resources and rural regions with limited infrastructure. This geographic and socioeconomic diversity tests the model’s adaptability to varying healthcare capabilities. Furthermore, Jiangsu’s epidemiologic representativeness—characterized by heterogeneous influenza transmission patterns and region-specific seasonal peaks (e.g., northern winter surges vs. southern bimodal outbreaks)—provides a robust natural experiment to evaluate model stability across fluctuating disease dynamics. This dual focus on clinical and epidemiologic variability ensures the model’s reliability in real-world settings, where both resource disparities and shifting transmission landscapes influence diagnostic accuracy.

#### 2.1.3 Trial Registration and Reporting Standards

The study was prospectively registered on the Open Science Framework (OSF) platform (DOI: 10.17605/OSF.IO/SC93Y) and the Chinese Clinical Trial Registry (ChiCTR2000028883), ensuring transparency and compliance with international clinical trial standards. Protocol development and reporting strictly adhere to the SPIRIT (Standard Protocol Items: Recommendations for Interventional Trials) 2013 guidelines (15), with a staged workflow encompassing model development, validation, and real-world deployment illustrated in **Figure 1**. This dual registration and standardized reporting framework enhances reproducibility, facilitates independent verification, and aligns with ethical and regulatory requirements for AI-integrated clinical research.

**Figure 1.**
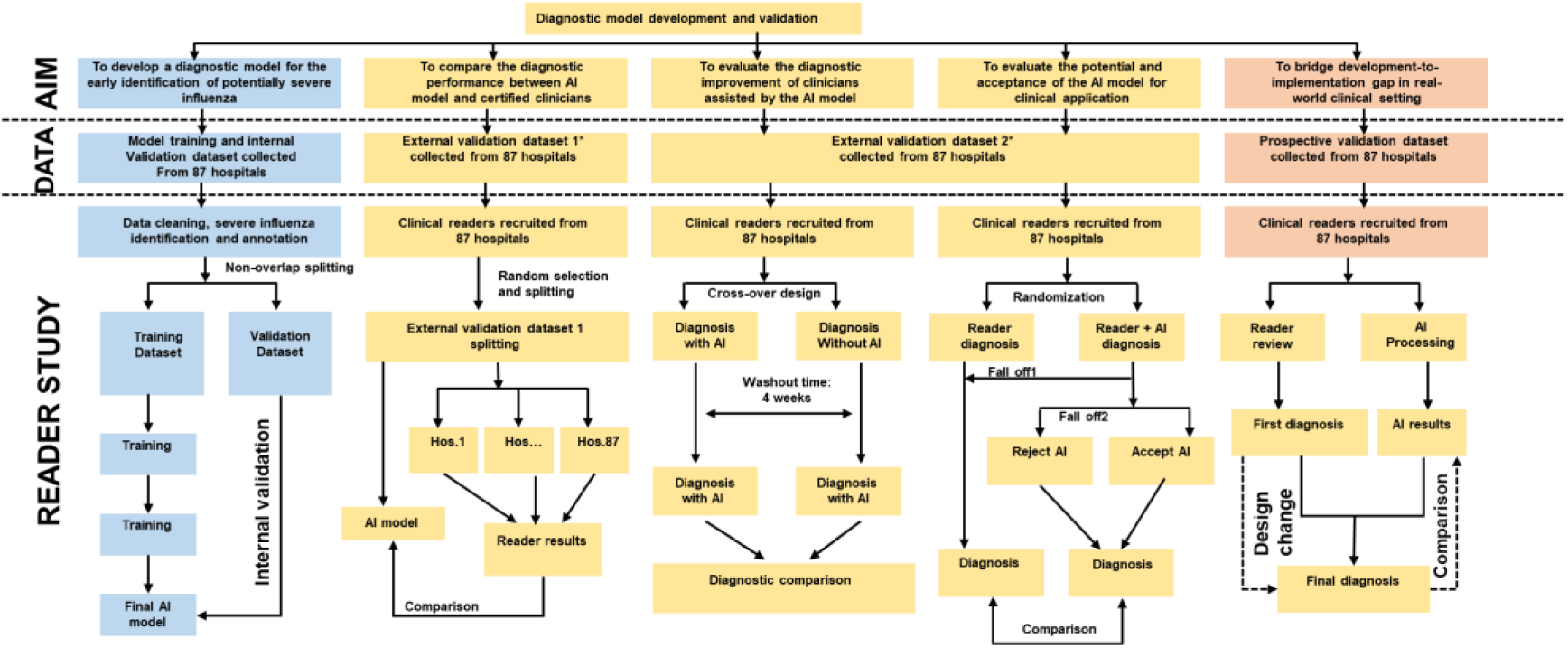
Study overview.

#### 2.1.4 Phase Overview

The study comprises five sequential phases, which is based on the multiple nesting approach used in Yan Bo’s past methods(16).

1. Model development: Retrospective data (2019–2024) from 87 hospitals for training and internal validation.
2. External validation-Comparative Study: Benchmarking model performance against clinician diagnoses.
3. Multi-reader, multi-case validation: Assessing inter-physician variability with/without model assistance.
4. Randomized controlled trial: Evaluating real-time diagnostic accuracy in a clinician-AI collaborative setting.
5. Prospective validation: Real-world testing during the 2025 influenza season.

#### 2.1.5 Key Modifications from Original Protocol

The original protocol was enhanced to strengthen methodological rigor and transparency: Explicit hospital inclusion criteria were introduced, including quantifiable thresholds such as minimum annual influenza case volume (≥100 confirmed cases) and standardized EHR documentation requirements, ensuring data quality and cross-hospital comparability.

The geographic rationale for selecting Jiangsu Province was clarified to address its heterogeneous healthcare infrastructure (urban vs. rural disparities) and diverse influenza epidemiology, providing a robust testbed for evaluating model generalizability.

To improve reporting transparency, adherence to the SPIRIT 2013 guidelines for clinical trial protocols was emphasized, and trial registration details (ChiCTR2000028883, OSF DOI: 10.17605/OSF.IO/SC93Y) were explicitly documented. These modifications ensure reproducibility, align with international standards, and address critical gaps in the original design.

### 2.2 Eligibility criteria

#### 2.2.1 Inclusion Criteria for Patients

1. Laboratory-confirmed influenza virus infection via nucleic acid testing (RT-PCR) or rapid antigen testing, adhering to China’s *Diagnosis and Treatment Protocol for Influenza*;
2. All age groups;
3. Written consent obtained from patients: (a) patients with full civil capacity (≥18 years); (b)Legal guardians for minors (<18 years), incapacitated adults, or patients with cognitive impairment (e.g., dementia).

#### 2.2.2 Exclusion Criteria for Patients

1. Co-infection with other respiratory pathogens (e.g., SARS-CoV-2, RSV) confirmed by multiplex PCR;
2. Critical illness unrelated to influenza at enrollment (e.g., trauma, sepsis);
3. (3)Incomplete baseline data (e.g., missing symptom onset date, laboratory results).

#### 2.2.3 Inclusion Criteria for Clinicians

1. ≥5 years of clinical experience in influenza diagnosis or participation in national-level influenza research projects (e.g., WHO-sponsored surveillance programs);
2. Willingness to complete protocol-mandated tasks, including dual-round diagnosis (baseline vs. model-assisted) and post-study interviews;
3. Currently practicing in a tertiary general hospital in Jiangsu Province.

#### 2.2.4 Exclusion Criteria for Clinicians

1. Temporary or part-time staff without direct patient care responsibilities;
2. Participation in conflicting AI-related clinical trials within the past 6 months.

### 2.3 Interventions

#### 2.3.1 Phase I: Model Development and Internal Validation

This phase utilizes retrospective electronic health records (EHRs) from 87 hospitals(**Table 1**) in Jiangsu Province (January 2019–December 2024), encompassing four key data categories:

1. Demographics: Age, sex, vaccination history, and comorbidities.
2. Clinical symptoms: Fever duration, cough frequency, dyspnea severity, and other symptoms detailed in Table 2.
3. Laboratory/imaging data: Viral subtype (confirmed via RT-PCR), blood cell counts, and chest X-ray/CT findings.
4. Outcomes: Severe influenza progression (defined as ICU admission or mechanical ventilation) and mortality.

**Table 1.**
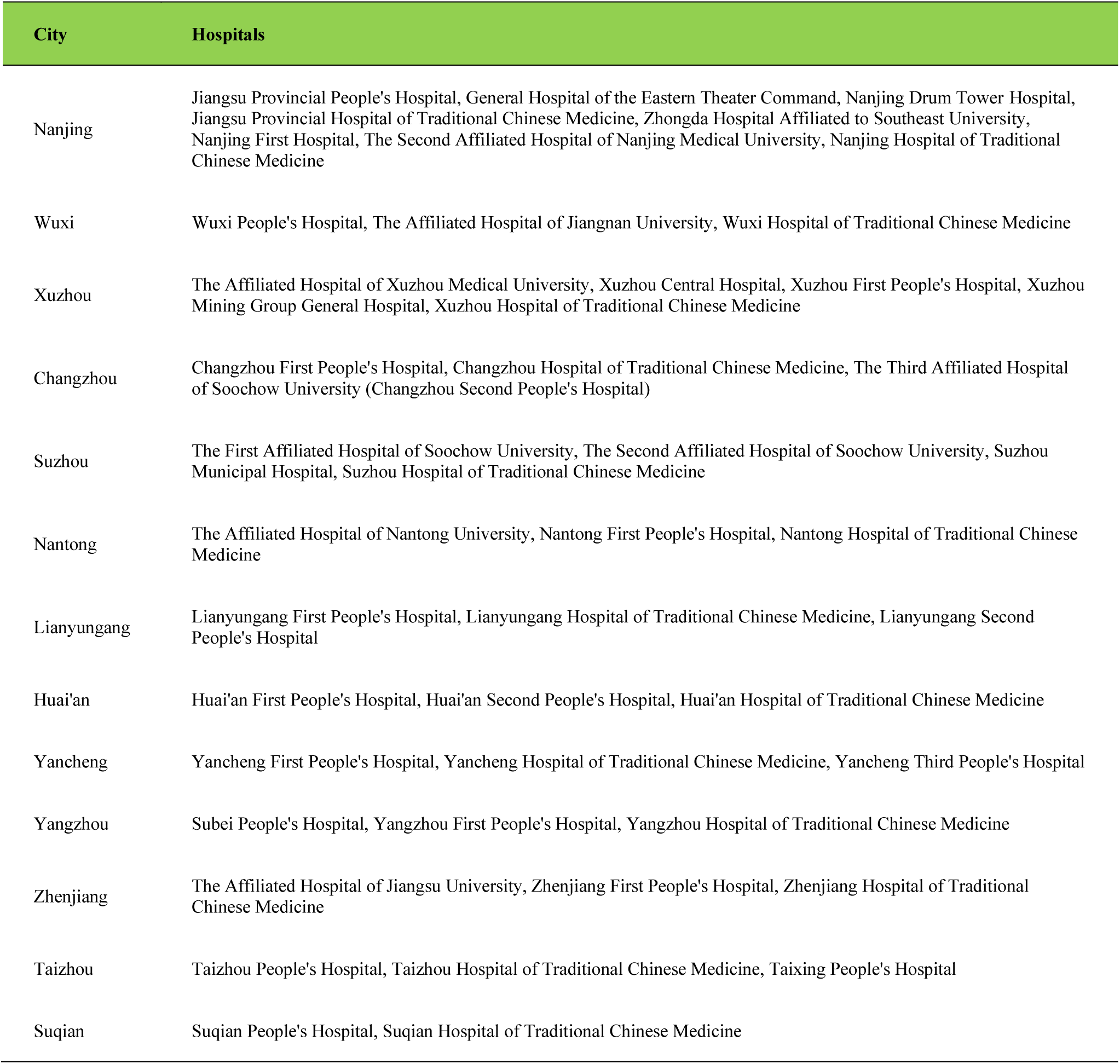
Datasets distribution.

| City | Hospitals |
| --- | --- |
| Nanjing | Jiangsu Provincial People's Hospital, General Hospital of the Eastern Theater Command, Nanjing Drum Tower Hospital, Jiangsu Provincial Hospital of Traditional Chinese Medicine, Zhongda Hospital Affiliated to Southeast University, Nanjing First Hospital, The Second Affiliated Hospital of Nanjing Medical University, Nanjing Hospital of Traditional Chinese Medicine |
| Wuxi | Wuxi People's Hospital, The Affiliated Hospital of Jiangnan University, Wuxi Hospital of Traditional Chinese Medicine |
| Xuzhou | The Affiliated Hospital of Xuzhou Medical University, Xuzhou Central Hospital, Xuzhou First People's Hospital, Xuzhou Mining Group General Hospital, Xuzhou Hospital of Traditional Chinese Medicine |
| Changzhou | Changzhou First People's Hospital, Changzhou Hospital of Traditional Chinese Medicine, The Third Affiliated Hospital of Soochow University (Changzhou Second People's Hospital) |
| Suzhou | The First Affiliated Hospital of Soochow University, The Second Affiliated Hospital of Soochow University, Suzhou Municipal Hospital, Suzhou Hospital of Traditional Chinese Medicine |
| Nantong | The Affiliated Hospital of Nantong University, Nantong First People's Hospital, Nantong Hospital of Traditional Chinese Medicine |
| Lianyungang | Lianyungang First People's Hospital, Lianyungang Hospital of Traditional Chinese Medicine, Lianyungang Second People's Hospital |
| Huai'an | Huai'an First People's Hospital, Huai'an Second People's Hospital, Huai'an Hospital of Traditional Chinese Medicine |
| Yancheng | Yancheng First People's Hospital, Yancheng Hospital of Traditional Chinese Medicine, Yancheng Third People's Hospital |
| Yangzhou | Subei People's Hospital, Yangzhou First People's Hospital, Yangzhou Hospital of Traditional Chinese Medicine |
| Zhenjiang | The Affiliated Hospital of Jiangsu University, Zhenjiang First People's Hospital, Zhenjiang Hospital of Traditional Chinese Medicine |
| Taizhou | Taizhou People's Hospital, Taizhou Hospital of Traditional Chinese Medicine, Taixing People's Hospital |
| Suqian | Suqian People's Hospital, Suqian Hospital of Traditional Chinese Medicine |

**Table 2.**
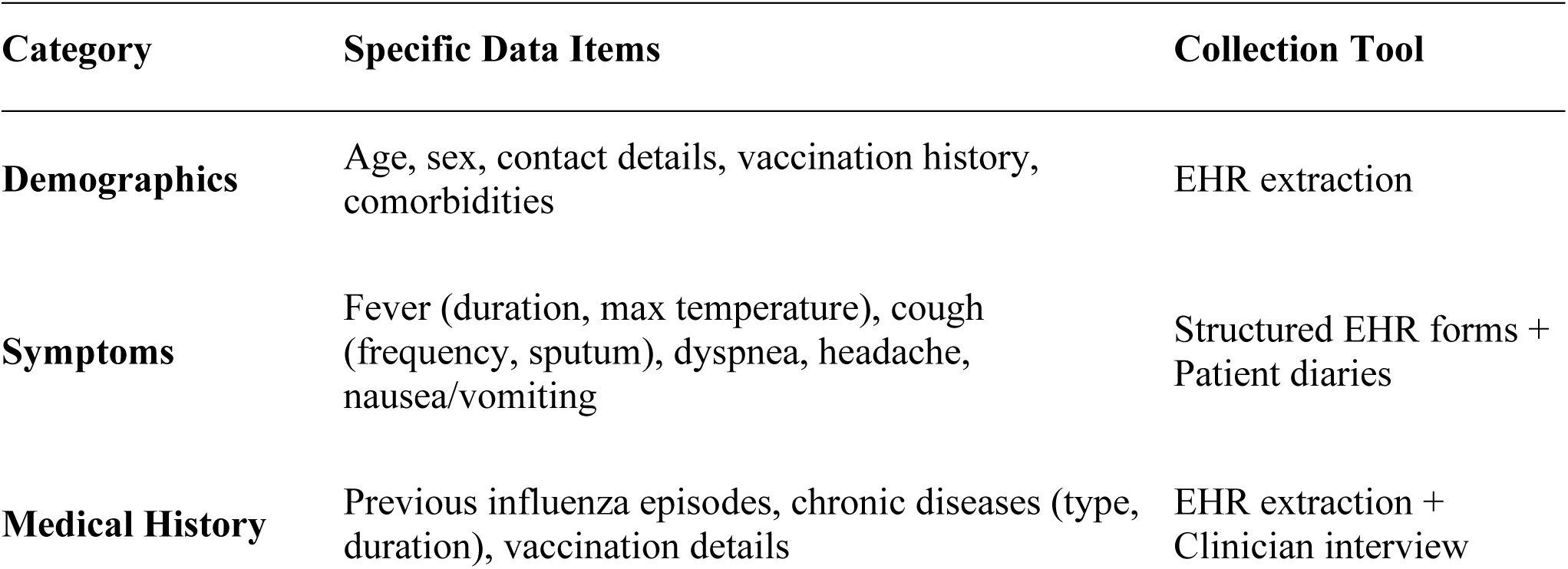
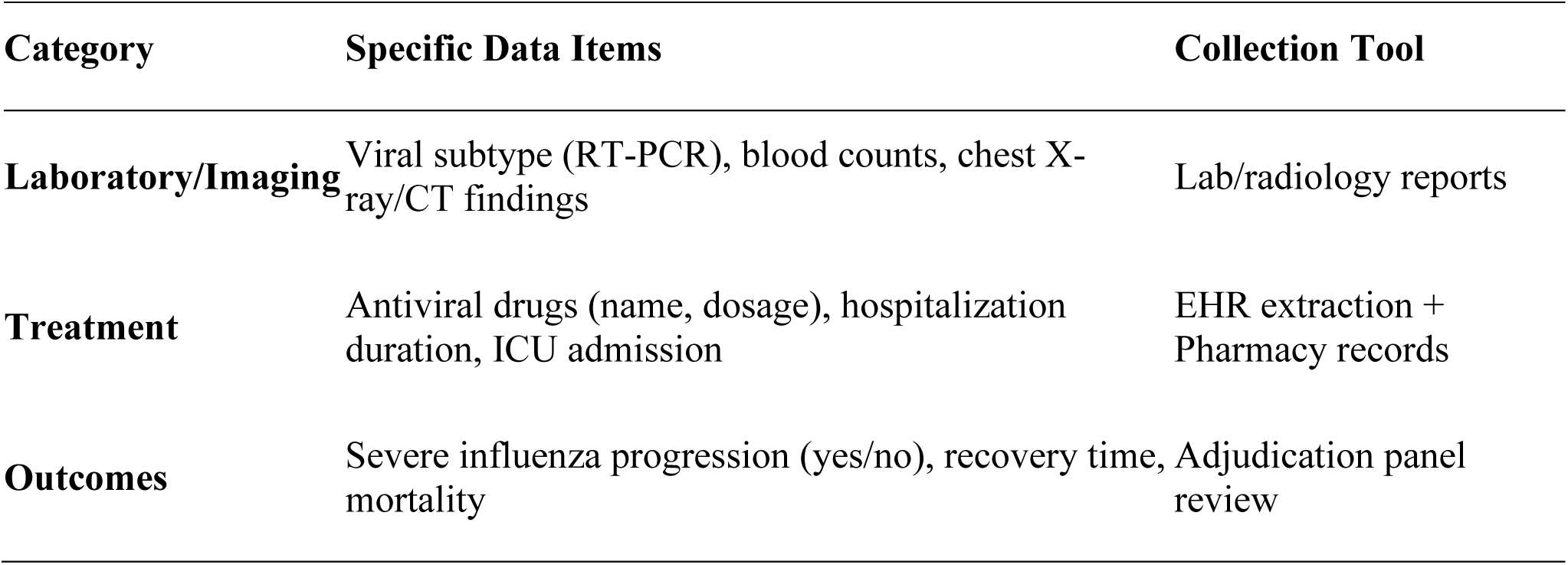
Data Collection Framework.

| Category | Specific Data Items | Collection Tool |
| --- | --- | --- |
| Demographics | Age, sex, contact details, vaccination history, comorbidities | EHR extraction |
| Symptoms | Fever (duration, max temperature), cough (frequency, sputum), dyspnea, headache, nausea/vomiting | Structured EHR forms + Patient diaries |
| Medical History | Previous influenza episodes, chronic diseases (type, duration), vaccination details | EHR extraction + Clinician interview |
| Laboratory/Imaging | Viral subtype (RT-PCR), blood counts, chest X-ray/CT findings | Lab/radiology reports |
| Treatment | Antiviral drugs (name, dosage), hospitalization duration, ICU admission | EHR extraction + Pharmacy records |
| Outcomes | Severe influenza progression (yes/no), recovery time, mortality | Adjudication panel review |

The dataset will be stratified by hospital and influenza season and partitioned into 70% training and 30% internal validation subsets, ensuring balanced representation of high-risk subgroups (e.g., infants, elderly). A ResNet-50-based deep neural network will be implemented using PyTorch, optimized via the AdamW algorithm (learning rate = 0.001, weight decay = 0.01) to minimize a weighted cross-entropy loss function, which assigns higher penalties to misclassifications of severe influenza cases to address class imbalance. This architecture integrates both structured clinical data and imaging features, enabling the model to learn complex patterns predictive of disease progression. Internal validation will assess baseline performance metrics (e.g., AUC, sensitivity) before advancing to external testing phases. ResNet-50 was adapted for structured clinical data by replacing convolutional layers with fully connected layers, retaining its residual learning framework to handle imbalanced outcomes.

#### 2.3.2 Phase II: External Validation-Comparative Study

This phase evaluates the model’s generalizability using 500 de-identified cases from external hospitals not included in Phase I. 218 physicians—stratified by experience level (resident, attending, chief)—will independently diagnose these cases without AI assistance, relying solely on clinical symptoms, lab results, and imaging findings. Concurrently, the same cases will be analyzed by the trained model to generate predictions. Diagnostic performance will be compared between clinicians and the model using metrics including AUC (area under the ROC curve), sensitivity, specificity, and time-to-diagnosis. This dual-assessment framework quantifies the model’s added value in real-world settings, benchmarking its accuracy and efficiency against human expertise while ensuring external validity through geographically and institutionally diverse case sources. Results will inform refinements before progressing to advanced validation phases.

#### 2.3.3 Phase III: Multi-Reader, Multi-Case Validation

This phase employs a dual-round diagnostic design to assess the impact of AI assistance on clinician performance. Each participating hospital will recruit 5–10 clinicians to independently evaluate cases in two sequential rounds:

1. Baseline diagnosis: Clinicians diagnose cases using standard protocols without AI input.
2. Model-assisted diagnosis: Clinicians review model-generated risk scores and key contributing factors (e.g., fever duration, lab abnormalities) before finalizing decisions.

This will be done to include every general hospital in every city in Jiangsu. All doctors had to do two rounds of diagnosis on the cases in the dataset(17). A review found that using a machine to help doctors make diagnoses can improve how well they do their job (18).To minimize bias, cases are randomly reordered between rounds, and a 2–4-week washout period separates the two evaluations to reduce recall effects. Diagnostic accuracy (measured by AUC) and clinician confidence (rated on a 5-point Likert scale) will be compared across rounds to quantify the model’s influence on decision-making consistency and practitioner certainty. This multi-reader, multi-case framework ensures robust evaluation of AI-human collaboration while addressing contextual variability in real-world clinical workflows.

#### 2.3.4 Phase IV: Randomized Controlled Trial

This phase employs a pragmatic randomized controlled design to evaluate the real-world efficacy of AI-assisted diagnosis. Clinicians (n=218) will be allocated 1:1 to either the control group (diagnosing cases using standard clinical judgment) or the model-assisted group (accessing AI predictions via a secure dashboard displaying risk scores and key contributing variables). Allocation will use block randomization stratified by hospital tier (urban vs. rural) and clinician experience level (resident, attending, chief), ensuring balanced representation across critical subgroups.

The control group will rely solely on conventional diagnostic workflows, while the model-assisted group will integrate AI outputs into their decision-making process, retaining full authority to accept or disregard recommendations. The primary outcome is the difference in AUC between groups, with a ΔAUC >0.15 predefined as clinically significant, aligning with FDA benchmarks for AI/ML-based diagnostic tools. This design mimics real-world clinical practice, quantifying the added value of AI in enhancing diagnostic accuracy while preserving clinician autonomy.

#### 2.3.5 Phase V: Prospective Validation Study

As Dai et al. illustrate the point, using prospective cohort data validation provides a better view of the model’s performance in the real world (19).This final phase evaluates the model’s real-world clinical utility during the 2025 influenza season. The AI model will be deployed in real time to analyze electronic health records (EHRs) of newly admitted influenza patients across all 87 hospitals.

Clinicians will receive model-generated predictions (e.g., risk scores, key contributing factors) within 2 hours of case entry via a secure, institution-specific dashboard.

Data collection will capture:

1. Clinician actions: Initial diagnosis (pre-model assessment), final diagnosis (post-model review), and time-to-decision (minutes from case entry to diagnosis).
2. Model performance: Concordance between model predictions and ground truth (severe influenza confirmed by an independent adjudication panel).

This prospective design validates the model’s operational feasibility, diagnostic timeliness, and accuracy in dynamic clinical environments, providing critical evidence for scalability and regulatory approval.

#### 2.3.6 Key Modifications from Original Protocol

The protocol was refined to address methodological gaps and enhance robustness: Technical specifications were expanded to detail the ResNet-50 architecture, optimization parameters (AdamW, learning rate = 0.001), and bias mitigation strategies (e.g., stratified sampling for high-risk subgroups). Stratified randomization was strengthened by implementing block randomization stratified by hospital tier (urban/rural) and clinician experience level (resident/attending/chief), ensuring balanced group allocation. Real-time workflow integration was clarified, specifying a 2-hour prediction turnaround for model outputs to align with clinical decision-making timelines. Finally, an independent adjudication panel was introduced to retrospectively validate severe influenza outcomes (e.g., ICU admission), reducing measurement bias and ensuring ground-truth reliability. These modifications collectively enhance the study’s rigor, transparency, and applicability to real-world healthcare settings.

### 2.4 Outcomes

#### 2.4.1 Primary Outcome

The Area Under the Receiver Operating Characteristic Curve (AUC) serves as the primary metric to evaluate the model’s diagnostic accuracy(20). The AUC quantifies the model’s ability to discriminate between patients who will progress to severe influenza and those who will not, integrating both sensitivity (true positive rate) and specificity (true negative rate). To account for variability in clinician interpretations across multi-reader studies, the Dorfman-Berbaum-Metz (DBM) method will be applied, which adjusts for inter-physician differences in case assessments (17). The use of this method allows for a more accurate assessment of the difference in AUC between the two groups by taking into account the variability in the diagnostic process between different physicians as readers (21). AUC values and their 95% confidence intervals will be calculated by constructing ROC curves using the MRMCaov (version 0.2.1) (22) and pROC (version 1.18.0) software packages (23). The 95% confidence intervals for these indicators will be calculated using the correlation method of binomial distribution, and the Clopper-Pearson method will be used for exact calculation (22). Differences between the model-assisted and control groups on these indicators will be compared by non-parametric tests of independent samples (e.g. Mann - Whitney U test) (24).Clinically significant improvement is defined as a ΔAUC >0.15 between the model-assisted and physician-only diagnostic groups, while the null hypothesis assumes no discriminative capacity (AUC ≤0.70, equivalent to random guessing). This threshold-based approach ensures statistical rigor while aligning with regulatory benchmarks for AI-driven diagnostic tools.

#### 2.4.2 Secondary Outcomes

Secondary endpoints include diagnostic sensitivity, defined as the proportion of true severe influenza cases correctly identified by the model or clinicians, which reflects the tool’s capacity to minimize missed diagnoses in high-risk populations (25). Diagnostic specificity, the proportion of non-severe cases accurately classified, aims to reduce unnecessary interventions for low-risk patients. Positive predictive value (PPV) quantifies the probability that a patient diagnosed as severe by the model or clinician will progress to severe disease, while negative predictive value (NPV) estimates the likelihood that a non-severe classification remains accurate. Additionally, the misdiagnosis rate— calculated as the average number of errors per 100 diagnoses—captures both false positives (non-severe cases incorrectly flagged as severe) and false negatives (severe cases erroneously classified as non-severe). These metrics collectively evaluate the model’s clinical utility, balancing accuracy with real-world diagnostic trade-offs.

#### 2.4.3 Subgroup Analyses

Predefined subgroup analyses will assess heterogeneity in model performance across key variables: patient characteristics—stratified by age (infants, children, adults, elderly), comorbidities (cardiovascular disease, diabetes), and vaccination status; clinician factors—including experience level (resident, attending, chief physician) and hospital resource tier (urban vs. rural facilities); and geographic variation—comparing northern versus southern regions of Jiangsu Province to evaluate the impact of local healthcare infrastructure and influenza epidemiology. These analyses aim to identify subgroups where the model excels or underperforms, informing targeted clinical deployment. Statistical adjustments, such as Bonferroni correction for multiple comparisons and multivariable regression for confounders (e.g., seasonal trends), will ensure robust interpretation of subgroup differences.

#### 2.4.4 Statistical Adjustment

To address potential biases, statistical adjustments were rigorously applied: multiple comparisons across predefined subgroups were controlled using the Bonferroni correction, where the significance threshold (α) was adjusted to *⍺* = 0.05/number of subgroups. Additionally, confounding variables— including seasonality, baseline viral load, and treatment delays—were adjusted via multivariable logistic regression, with these factors modeled as covariates to isolate the independent effect of the AI model on diagnostic accuracy. These adjustments ensure robust interpretation of subgroup differences and mitigate inflation of type I error rates or spurious associations, enhancing the validity and reliability of study findings.

#### 2.4.5 Handling Missing Data

To address missing data, variables with >20% missing values (e.g., incomplete vaccination records) will be excluded to ensure reliability. For remaining gaps, the Multivariate Imputation by Chained Equations (MICE) package in R will be employed, leveraging iterative modeling across variables to impute missing values under the missing-at-random assumption. This method preserves dataset integrity by estimating missing entries based on observed patterns, thereby minimizing bias while maintaining statistical power.

#### 2.4.6 Adjudication of Ground Truth

To ensure accurate outcome classification, an independent adjudication panel comprising three senior infectious disease specialists will retrospectively review all cases classified as severe influenza, verifying endpoints such as ICU admission, mechanical ventilation, or mortality. Discrepancies in outcome assessments among panel members will be resolved through majority vote, with detailed documentation of dissenting opinions. This rigorous process minimizes misclassification bias and enhances the reliability of outcome data, serving as the gold standard for model and clinician performance evaluation.

#### 2.4.7 Key Modifications from Original Protocol

The protocol was refined to enhance methodological rigor and transparency: AUC interpretation was clarified by defining explicit thresholds for clinical significance (ΔAUC >0.15) and null hypothesis testing (AUC ≤0.70, equivalent to random guessing), ensuring alignment with regulatory benchmarks. Subgroup analyses were strengthened through predefined statistical adjustments, including Bonferroni correction for multiple comparisons and multivariable regression to control for confounders such as seasonal trends. A robust missing data protocol was formalized, specifying exclusion criteria for variables with >20% missingness and adopting the MICE (Multivariate Imputation by Chained Equations) methodology to impute remaining gaps under missing-at-random assumptions. Finally, an independent adjudication panel was introduced to retrospectively validate severe influenza outcomes (e.g., ICU admission, mortality) via majority consensus, minimizing misclassification bias. These modifications collectively address limitations in the original design, improving validity, reproducibility, and ethical compliance.

### 2.5 Sample size

#### 2.5.1 Calculation Rationale

The sample size was determined based on the primary outcome (AUC comparison between model-assisted and clinician-only diagnoses) using a two-group superiority design. Key parameters were derived from prior studies on AI diagnostic tools (26,27):

Expected effect size: ΔAUC = 0.15 (clinically meaningful improvement).

Type I error (α): 0.05 (two-tailed).

Power (1-β): 0.95.

Attrition rate: 20% adjustment for patient dropout or incomplete data.

The minimum sample size for AUC comparison was calculated using the following formula(28):

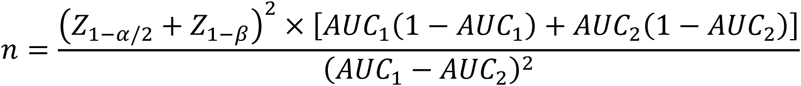

Where:

*AUC*_1_ (model-assisted): 0.85 (based on Yang et al., 2023 (29)).

*AUC*_2_ (clinician-only): 0.70 (null hypothesis).

*Z*_1−*⍺*/2_= 1.96, *Z*_1−*β*_= 1.645.

#### 2.5.2 Sample Size Derivation

The sample size calculation began with an initial estimate of 145 patients per group (290 total), derived from a power analysis targeting a clinically significant ΔAUC of 0.15 (α=0.05, β=0.95). To account for potential 20% attrition due to incomplete data or participant dropout, the final patient sample was adjusted to 174 per group (348 total). For clinicians, a minimum of 218 participants (109 per group) was required to ensure balanced representation across experience levels (resident, attending, chief) while addressing hospital-level clustering effects, as recommended by methodology for cluster-randomized trials (30). This dual-tiered approach balances statistical precision with pragmatic considerations, ensuring robust evaluation of the AI model’s impact in diverse clinical environments.

#### 2.5.3 Final Allocation

The study will enroll a total of 348 patients, comprising both retrospective cohorts (historical data from 2019–2024) and prospective cohorts (real-time data from the 2025 influenza season). Clinician participation includes 218 practitioners, stratified by hospital tier (urban vs. rural) and seniority level (resident, attending, or chief physician) to ensure balanced representation of clinical expertise and institutional resource variability. This allocation strategy optimizes statistical power while reflecting real-world healthcare diversity, supporting robust validation of the AI model across heterogeneous settings.

#### 2.5.4 Justification

The patient sample size was determined through a power analysis to ensure ≥80% statistical power for detecting a clinically meaningful improvement (ΔAUC >0.15) in diagnostic accuracy, aligning with FDA guidelines for validating AI-driven medical devices (31). The clinician sample was designed to reflect real-world diversity in clinical expertise (e.g., residents to chief physicians) and practice settings (urban vs. rural hospitals), ensuring generalizability across heterogeneous healthcare environments. This dual focus on statistical rigor and ecological validity balances methodological precision with practical applicability, addressing both regulatory standards and real-world implementation challenges.

#### 2.5.5 Key Modifications from Original Protocol

Critical refinements were made to strengthen methodological rigor. Formula transparency was enhanced by explicitly defining the AUC-based sample size calculation formula (ΔAUC >0.15, α=0.05, β=0.95) and its parameters, ensuring reproducibility. Attrition adjustment incorporated a 20% buffer to account for potential data loss or participant dropout, improving robustness in real-world settings. Stratification protocols were clarified to allocate clinicians by both experience level (resident, attending, chief) and hospital tier (urban vs. rural), ensuring balanced representation across subgroups. Finally, regulatory alignment was achieved by citing FDA guidelines for AI/ML-based diagnostics to justify the predefined effect size thresholds, reinforcing clinical relevance and compliance with regulatory standards(31). These modifications collectively address limitations in the original protocol, enhancing validity and transparency.

### 2.6 Assignment of interventions

#### 2.6.1 Randomization and Allocation

A computer-generated randomization sequence will be created using the blockrand package in R, stratified by hospital tier (urban vs. rural) and clinician experience level (resident, attending, or chief physician). This approach ensures balanced allocation across subgroups, minimizing confounding effects from institutional resources or practitioner expertise. Participants will be assigned to either the control group (standard clinical diagnosis) or the model-assisted group (AI-supported diagnosis) at a 1:1 allocation ratio. The randomization sequence will be concealed in sequentially numbered, opaque envelopes managed by an independent statistician to prevent selection bias, ensuring equitable distribution of participants and maintaining trial integrity.

To ensure unbiased group assignment, an independent statistician uninvolved in recruitment or data analysis will prepare opaque, sequentially numbered envelopes containing group allocations (control or model-assisted). These sealed envelopes will be securely stored and only opened sequentially by a dedicated study coordinator after clinicians complete baseline assessments. This procedure rigorously prevents selection bias by concealing allocation details until the point of intervention assignment, thereby safeguarding the randomization integrity and ensuring equitable distribution of participants across study groups.

#### 2.6.2 Blinding (Masking)

To minimize bias, a single-blind design will be implemented for clinicians: those in the control group will remain unaware of their allocation status and the existence of the AI model, while clinicians in the model-assisted group will access predictions through a neutral interface labeled as a “Decision Support Tool,” avoiding explicit references to AI. Patients will be fully blinded to group assignments, with informed consent documents describing participation in “a study to improve influenza diagnosis” without disclosing AI involvement. Independent outcome adjudicators assessing severe influenza outcomes will also remain blinded to group allocations and model outputs, ensuring objective evaluation of clinical endpoints. This multi-layered blinding strategy safeguards against performance and detection bias, maintaining the trial’s scientific rigor.

#### 2.6.3 Intervention Groups

In the control group, clinicians will diagnose influenza cases using standard clinical protocols—integrating symptoms, laboratory results, and imaging findings—without any AI assistance. Patients in this group will receive routine care based solely on clinician assessments. In contrast, the model-assisted group will utilize a secure digital dashboard providing real-time AI predictions, including a risk score (0–100%) for severe influenza progression and highlighted contributing factors (e.g., prolonged fever duration, abnormal lymphocyte counts). While the model offers data-driven insights, its outputs are non-prescriptive; clinicians retain full authority to accept, modify, or reject recommendations, ensuring clinical judgment remains central to decision-making. This design balances technological support with practitioner autonomy, reflecting real-world diagnostic workflows.

#### 2.6.4 Handling Cross-Over and Contamination

To preserve the integrity of group assignments, cross-over between groups is strictly prohibited: clinicians allocated to the control group will be denied access to the AI model throughout the trial. To mitigate contamination risks, separate electronic health record (EHR) access permissions will be enforced for control and model-assisted groups, ensuring clinicians cannot inadvertently view or utilize AI outputs outside their assigned protocol. Additionally, regular audits (monthly reviews of EHR access logs and diagnostic workflows) will be conducted to verify compliance with allocation protocols. These measures collectively safeguard against data leakage and intervention overlap, maintaining the study’s internal validity.

#### 2.6.5 Ethical Considerations

All participants will provide informed consent: patients or their legal guardians will consent to the use of their anonymized clinical data while remaining unaware of group assignments, with study participation described broadly as “research to improve influenza diagnosis.” Clinicians will consent to randomization but remain blinded to allocation mechanics to prevent bias. Emergency unblinding will be permitted only in cases where a clinician identifies a critical patient safety concern (e.g., life-threatening diagnostic uncertainty) requiring full transparency of diagnostic data. This exception will be documented and reviewed by the ethics committee to ensure compliance with ethical standards. These protocols balance participant autonomy, clinical safety, and methodological rigor, aligning with international guidelines for AI-integrated clinical trials.

#### 2.6.6 Key Modifications from Original Protocol

The protocol was refined to enhance methodological rigor and ethical compliance: stratified randomization now incorporates hospital tier (urban vs. rural) and clinician experience level as key variables, improving balance across subgroups. A detailed blinding protocol specifies single-blind procedures for clinicians (masked to allocation mechanics) and full blinding for patients and outcome adjudicators. To prevent contamination, technical safeguards—including tiered EHR access permissions—were implemented to isolate data flows between control and model-assisted groups. Lastly, ethical transparency was strengthened by defining explicit criteria for emergency unblinding (e.g., critical safety risks), ensuring alignment with Good Clinical Practice (GCP) guidelines. These revisions collectively address potential biases and operational challenges identified in the original design.

### 2.7 Data collection

#### 2.7.1 Data Sources and Tools

The study leverages electronic health records (EHRs) from 87 hospitals in Jiangsu Province, China, spanning retrospective (January 2019–December 2024) and prospective (January–December 2025) periods. Standardized EHR fields include demographics, symptoms, laboratory results, imaging reports, treatment details, and outcomes (**Table 2**), with data completeness ensured through automated validation checks (e.g., alerts for missing values) and manual audits of 5% randomly selected records. Clinician diagnostic workflows are captured via customized digital forms documenting baseline and model-assisted assessments, alongside time-stamped logs tracking diagnostic duration and patterns of AI tool interaction (e.g., frequency of accessing risk scores). Patient-reported outcomes are supplemented by optional daily symptom diaries administered through a WeChat mini-program, enabling real-time symptom tracking for consenting participants. This multi-modal data framework balances granular clinical detail with patient-centric insights, ensuring robust validation of the AI model across diverse data streams.

#### 2.7.2 Data Collection Timeline

The study’s data collection is structured into two distinct phases: retrospective (Phases I–II) and prospective (Phases III–V). During the retrospective phase, electronic health record (EHR) data spanning 2019 to 2024 will be extracted from all 87 participating hospitals via institution-specific application programming interfaces (APIs), with completion targeted within six months to ensure timely progression to subsequent stages. In the prospective phase, real-time data collection will occur throughout the 2025 influenza season, capturing clinical, diagnostic, and outcome variables as they emerge. These data will be synchronized daily to a centralized, secure database to maintain continuity and enable immediate analysis. This phased approach balances historical insights with dynamic, real-world validation, ensuring comprehensive evaluation of the AI model across diverse temporal and operational contexts.

#### 2.7.3 Quality Control Measures

To ensure data accuracy and consistency, comprehensive quality control protocols were implemented. Standardized training was provided to clinicians and data entry staff, focusing on EHR documentation practices (e.g., symptom coding conventions and structured data entry). Automated validation rules were applied to detect anomalies, including range checks (e.g., body temperature ≤ 42°C) and logical consistency validations (e.g., ensuring symptom onset dates precede diagnosis dates). Additionally, manual audits were conducted by independent reviewers, who cross-validated 10% of randomly selected records against source documents (e.g., laboratory reports, imaging files) to verify data fidelity. These layered measures—combining education, technology, and human oversight—minimize errors and enhance the reliability of the dataset for robust model validation.

#### 2.7.4 Missing Data Handling

To minimize missing data, mandatory fields (e.g., viral subtype, symptom onset date) are enforced in digital entry forms, ensuring critical variables are consistently documented. For remaining missing values, multiple imputation via chained equations (MICE package) will be applied to continuous variables, leveraging iterative regression models to estimate plausible values under missing-at-random assumptions. Categorical variables with <10% missingness will undergo mode imputation, replacing gaps with the most frequent category, while variables exceeding this threshold will be excluded to avoid bias from unreliable estimations. This tiered approach balances data completeness with methodological rigor, preserving statistical validity while addressing real-world data imperfections.

#### 2.7.5 Data Storage and Security

To safeguard participant confidentiality, all patient identifiers (e.g., names, contact information) will be removed and replaced with unique, non-traceable study IDs. Anonymized data will be stored in encrypted formats on password-protected servers hosted through the Open Science Framework (OSF) platform (osf.io/ayj75), a secure repository compliant with international data protection standards. Access to the dataset will be restricted to authorized researchers via two-factor authentication, ensuring that only verified personnel with explicit permissions can retrieve or modify the data. This multi-layered security framework aligns with GDPR and China’s Data Security Law, prioritizing privacy while enabling rigorous scientific analysis.

#### 2.7.6 Key Modifications from Original Protocol

The protocol was revised to enhance data integrity and compliance: Enhanced standardization introduced mandatory fields (e.g., symptom onset date) and validation rules (e.g., logical consistency checks) to ensure uniformity across multi-center EHR entries. Patient-reported tools were expanded to include WeChat-based symptom diaries, leveraging the platform’s widespread adoption in China for real-time, longitudinal tracking of patient-reported outcomes. Quality control protocols were strengthened by specifying automated validation algorithms (e.g., outlier detection) alongside manual audits (10% random record verification) to minimize data entry errors. Finally, transparency measures were prioritized through detailed documentation of data anonymization (e.g., unique study IDs) and encrypted storage on GDPR- and China’s Data Security Law-compliant platforms (OSF), ensuring ethical and legal adherence while maintaining research reproducibility. These refinements collectively address gaps in the original design, fostering rigor, participant engagement, and regulatory alignment.

### 2.8 Statistical analysis

#### 2.8.1 Primary Outcome Analysis

The Area Under the Curve (AUC) will serve as the primary metric to evaluate the diagnostic accuracy of the AI model. To account for variability inherent in multi-reader, multi-case study designs, the Dorfman-Berbaum-Metz (DBM) method will be employed, which adjusts for differences in clinician interpretations and case complexity. Analyses will be conducted using the MRMCaov (v0.2.1) and pROC (v1.18.0) packages in R, generating AUC values with 95% confidence intervals (CIs) derived from non-parametric bootstrapping (1,000 iterations). A clinically significant improvement is predefined as a ΔAUC >0.15 between the model-assisted and control groups, with the null hypothesis assuming no discriminative capacity (AUC ≤0.70, equivalent to random guessing). This approach ensures robust statistical inference while addressing real-world diagnostic variability across clinicians and settings. Hyperparameter optimization was conducted via Bayesian optimization with 5-fold cross-validation, prioritizing AUC on the validation set.

#### 2.8.2 Secondary Outcomes Analysis

Secondary endpoints include diagnostic sensitivity and specificity, calculated using the Clopper-Pearson exact method for binomial proportions to determine the model’s and clinicians’ ability to correctly identify true positive and true negative cases, respectively. Differences between clinician and model performance will be assessed via the Mann-Whitney U test, a non-parametric method suitable for skewed distributions. Positive and negative predictive values (PPV/NPV) will be adjusted for influenza prevalence using Bayes’ theorem and reported with 95% confidence intervals to reflect their clinical utility in real-world settings. The misdiagnosis rate, defined as the sum of false positives and false negatives per 100 diagnoses, will be analyzed using an independent samples t-test (if normally distributed) or Wilcoxon rank-sum test (for non-normal data), quantifying the model’s impact on reducing diagnostic errors. These metrics collectively evaluate the model’s precision, reliability, and practical value in augmenting clinical decision-making.

#### 2.8.3 Subgroup Analyses

Subgroup analyses will evaluate the model’s performance heterogeneity across predefined categories: patient-level factors (age [infants, children, adults, elderly], comorbidities, and vaccination status), clinician-level factors (experience [resident, attending, chief] and hospital resource tier [urban vs. rural]), and geographic variation (northern vs. southern Jiangsu Province). To mitigate bias, statistical adjustments include Bonferroni correction for multiple comparisons (adjusted α = 0.05 / number of subgroups) and multivariable logistic regression controlling for confounders such as seasonal trends, baseline viral load, and treatment delays. These analyses aim to identify subgroups where the model excels or underperforms, ensuring tailored clinical deployment and validating robustness across diverse healthcare settings.

#### 2.8.4 Missing Data Handling

To address missing data, variables with >20% missing values (e.g., incomplete vaccination records) will be excluded to maintain analytical reliability. For remaining gaps, multiple imputation via chained equations (MICE package in R) will generate 10 imputed datasets for continuous variables, preserving statistical power under missing-at-random assumptions. Categorical variables with <10% missingness will undergo mode imputation, replacing missing entries with the most frequent category, while those exceeding this threshold will be excluded to avoid biased estimations. This protocol balances data completeness with methodological rigor, minimizing potential distortions in model performance evaluation.

#### 2.8.5 Sensitivity Analyses

To evaluate the robustness of findings, sensitivity analyses will include complete case analysis (comparing results with and without imputation) and model stability assessments (quantifying AUC variation across imputed datasets). These analyses ensure conclusions are not unduly influenced by missing data assumptions. For computational workflows, R (v4.3.1), Python (v3.10), and STATA (v18) will be utilized, integrating specialized packages for imputation (MICE), AUC calculation (pROC), and regression modeling. Reporting will adhere to the TRIPOD guidelines for transparent validation of predictive models, ensuring methodological transparency, reproducibility, and alignment with international standards for AI-driven diagnostic research.

#### 2.8.6 Key Modifications from Original Protocol

To enhance methodological rigor, the protocol was updated as follows: Inter-reader variability in AUC calculation was explicitly addressed using the Dorfman-Berbaum-Metz (DBM) method, which accounts for differences in clinician interpretations across multi-reader studies. Parametric assumptions for confidence intervals (CIs) were replaced with non-parametric bootstrapping (1,000 iterations) to improve robustness and reduce reliance on distributional assumptions. Sensitivity analyses were expanded to include model stability assessments, evaluating AUC consistency across multiple imputed datasets to ensure results are not skewed by imputation methods. For subgroup analyses, Bonferroni correction thresholds (adjusted α = 0.05/number of subgroups) and multivariable regression models were formalized to adjust for confounding variables (e.g., seasonal trends, treatment delays). These modifications collectively strengthen the study’s validity, transparency, and alignment with best practices in AI-driven diagnostic research.

## 3 Ethics and dissemination

This study received ethical approval from the Ethics Committee of the Second Affiliated Hospital of Soochow University (Approval No. JD-LK-2019-106-01) and The Affiliated Hospital of Yangzhou University (Approval No. 2024-10-02). Annual renewals of ethical approval will be submitted to ensure ongoing compliance with evolving regulatory and ethical standards. Informed consent was obtained from all participants: patients or their legal guardians (for minors or incapacitated adults) provided written consent for anonymized data collection and analysis, while clinicians signed agreements acknowledging their roles in diagnosis and model interaction, with the explicit right to withdraw at any time. To safeguard privacy, all patient identifiers (e.g., names, contact details) were replaced with unique study codes, and anonymized data were stored on the Open Science Framework (OSF) platform (osf.io/ayj75), accessible only to authorized researchers via two-factor authentication. These measures align with international data protection standards (e.g., GDPR) and China’s Data Security Law, ensuring ethical integrity and participant confidentiality throughout the study.

### 3.1 Risk Mitigation

To ensure patient safety and research integrity, clinicians retain full authority to override model recommendations if conflicting with clinical judgment, while an independent adjudication panel reviews all severe outcomes (e.g., mortality) to confirm diagnostic accuracy. Emergency unblinding is permitted only when critical safety concerns necessitate full diagnostic transparency, with such instances documented and reviewed by ethics committees.

### 3.2 Dissemination Plan

For broader dissemination, results will be shared across all 87 participating hospitals via workshops and a bilingual Clinical Decision Support Guide, with plans to integrate model outputs into EHR systems post-regulatory approval. Findings will be submitted to journals (e.g., Frontiers in Medicine) and presented at international forums (e.g., WHO Global Influenza Meetings), complemented by public health bulletins highlighting implications for high-risk populations.

### 3.3 Data Sharing Policy

De-identified datasets and analysis code will be publicly accessible on the OSF platform post-publication, adhering to GDPR and China’s Data Security Law, while sensitive data (e.g., clinician interaction logs) require steering committee approval.

### 3.4 Key Modifications from Original Protocol

Key protocol modifications include enhanced data security (two-factor authentication, encrypted storage), annual ethical reviews, tiered data accessibility, and full transparency in model and clinician interaction protocols. These measures collectively prioritize ethical rigor, clinical relevance, and global scalability of the AI-driven diagnostic solution.

## Supporting information

Chictr20250113214030

## 6 Acknowledgments

This is a registered study (ChiCTR2000028883). The Ethics Committee of the Second Affiliated Hospital of Soochow University (JD-LK-2019-106-01) and the Affiliated Hospital of Yangzhou University (2024-10-02) approved this study.

Registration DOI: https://doi.org/10.17605/OSF.IO/SC93Y

## 7 Data Availability Statement

This is a research agreement for a clinical trial. Any information or data collected during the study will be stored on the OSF platform (osf.io/ayj75).

